# Linking physical food outlets to online platforms: A cross-sectional machine learning approach to analysing socioeconomic variations in Great Britain

**DOI:** 10.1101/2025.03.06.25323505

**Authors:** Jody C Hoenink, Yuru Huang, Jean Adams

## Abstract

**Objectives:** Physical food outlets are increasingly offering delivery through Online Food Delivery Service (OFDS) platforms, but the scale of this expansion remains unclear due to the labour-intensive process of manually matching outlets to online platforms. Understanding the share of outlets offering delivery is important, as it impacts food availability and thus potentially influences dietary behaviours. This paper demonstrates how a machine learning model can efficiently match physical to online outlets. We also analysed how the proportion of physical outlets listed online and online-only outlets varies by area-level deprivation.

**Methods:** The physical locations of outlets selling food in Great Britain was obtained from a centrally held register for food hygiene data, while online outlet data was collected through web scraping an OFDS platform. We calculated string distances based on outlet names and postcodes, which were then used to train a Random Forest model to match outlets from the two lists. Area-level deprivation was assessed using the Index of Multiple Deprivation.

**Results:** The Random Forest classifier model achieved an F1 score of 90%, a recall of 98%, and a precision of 83%. Overall, the median percentage of physical outlets also listed online was 14% (IQR 0 - 23), and the median percentage of online-only outlets was also 14% (IQR 0 - 27). The proportion of physical outlets listed online and online-only outlets was highest in more deprived areas. For example, compared to the least deprived areas, the most deprived areas were associated with a 6% greater proportion of physical food outlets listed online (95%CI 5%–6%) and a 3% greater proportion of online-only outlets (95%CI 1%–4%).

**Conclusion:** This study demonstrates the potential of machine learning techniques to efficiently match physical and online food outlets. This automated approach can provide insights into the relationship between physical and online food availability. Researchers and policymakers can use this method to better understand inequalities in food outlet availability and monitor the expansion of online delivery services.

## Introduction

The expansion of Online Food Delivery Service (OFDS) platforms such as Just Eat and Uber Eats has increased the accessibility of food prepared out-of-home (1). These platforms connect individuals with a broad range of food outlets, from local restaurants to major fast-food chains. In addition to increasing accessibility to foods from physical food outlets, OFDS platforms have also enabled the growth of online-only food outlets (i.e. dark kitchens). The rise of these platforms presents new challenges for understanding inequalities in food accessibility across socioeconomic areas, especially when trying to compare traditional physical food environments to the newer online space.

Previous research evaluated both the number of physical and online food outlets, as well as the ratio between them (2). However, the lack of linkage between physical and online outlets prevented the accurate estimation of the proportion of physical outlets that were also registered online or, conversely, online outlets with a physical presence. The scale of such a linkage was likely a barrier given the need to connect 79,000 online outlets from one OFDS platform (1) to a database containing over 620,000 physical food outlets (3).

Linking physical and online food outlets would facilitate more accurate exposure assessments and allow for ongoing monitoring of the food environment, such as identifying which outlets are more likely to have an online presence and exploring variations by socioeconomic position. This study demonstrates a proof of concept by applying machine learning to link datasets from a central food hygiene register with those on popular OFDS platforms in Great Britain (GB), assessing how physical and online-only outlets varied across area-level deprivation.

## Methods

Physical outlet locations were sourced from the Food Standards Agency (FSA)’s hygiene rating scheme data and matched with online outlet data obtained from the OFDS platform Just Eat Takeaway.com (Just Eat) using machine learning. Figure 1 describes the process by which we sourced data to match physical and online food outlets. Cross-sectional analyses were performed at the Middle Layer Super Output Area (MSOA) level, which are geographic units comprising 2,000–6,000 households. The study adhered to the UK Office for National Statistics’ web scraping policy (4) and used publicly available data. Thus, ethical approval was not required.

**Figure 1.**
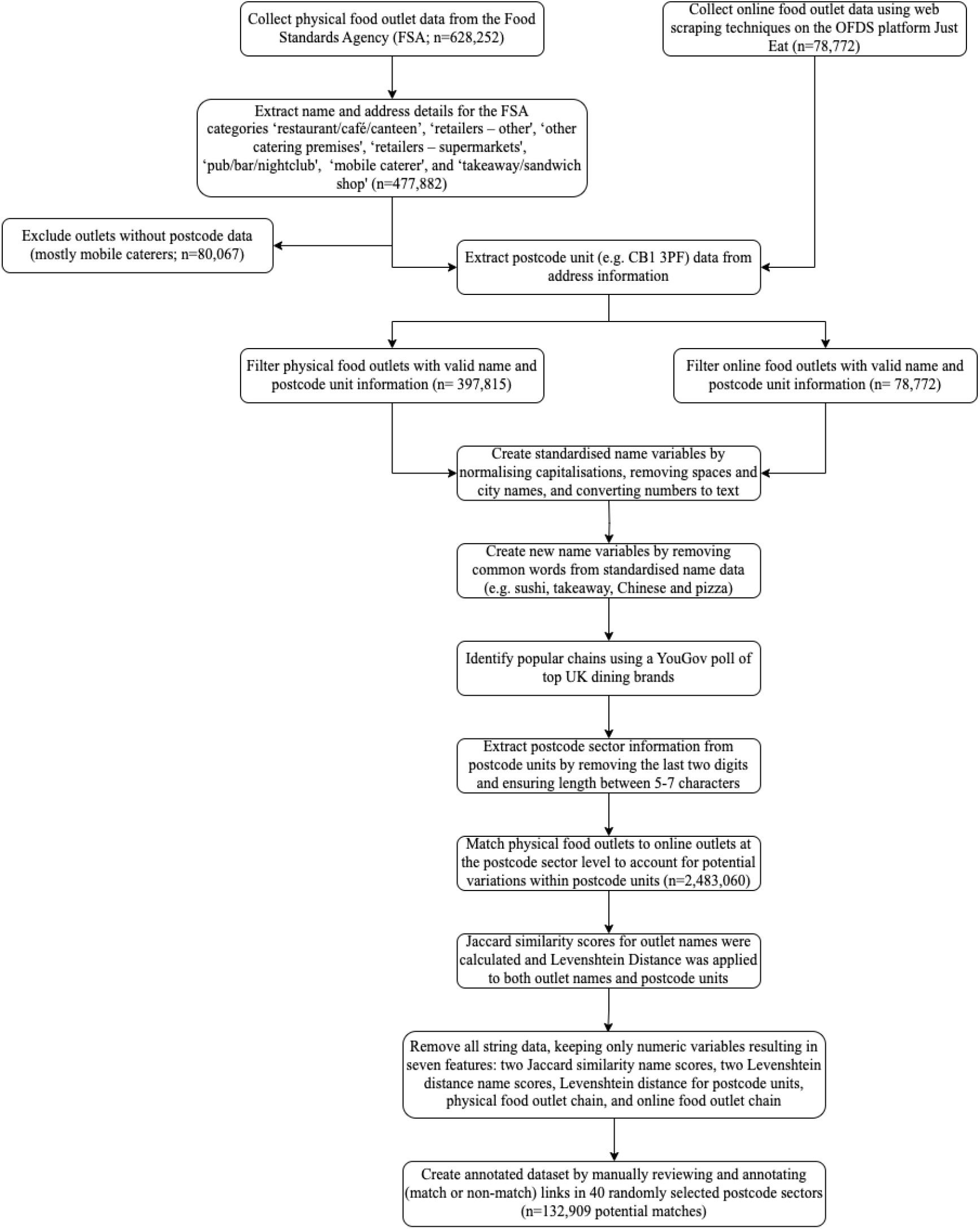
Flow of steps used to prepare data for the machine learning approach matching physical and online food outlets using postcode data and similarity metrics

### Data sources

#### Food Standards Agency

To compile a list of physical food outlets across GB, food hygiene data from FSA as of March 2024 was used (3). Information on the categories of food outlets, such as restaurants and supermarkets, was extracted directly from this dataset. Previous research has shown that FSA data closely aligns with street audits (5). Outlets without postcode information, primarily those categorised as mobile caterers, were excluded (16%; Figure 1).

#### OFDS platform

According to Statista, in 2022, Just Eat was the most popular OFDS platform in the United Kingdom, with twice as many users as the next most commonly used platform, Uber Eats (6). Similar to our previous study (7), the names and addresses of food outlets on Just Eat delivering to postcode sectors in GB were scraped using a web crawler developed in Python Scrapy framework. Data were collected over three days in March 2024.

### Physical and online food outlets

We included two outcome measures. First, we measured the proportion of physical outlets also listed online, calculated as the number of physical food outlets matched to an online outlet divided by the total number of physical food outlets at MSOA level. Second, we measured the proportion of online-only outlets, calculated as 1 minus the number of online food outlets matched to a physical outlet divided by the total number of online food outlets at MSOA level.

To facilitate the matching of physical and online food outlets, outlet names in both datasets were standardised and common terms like ‘takeaway’ were removed (Figure 1). We developed a third variable indicating whether an outlet was part of a popular chain or independent, using a YouGov poll of the most popular UK dining brands, an approach used by Kalbus et al. (8). After data cleaning, physical outlets were linked to online outlets at the postcode sector (e.g. CB1 3) level instead of the full postcode (e.g. CB1 3PF) to account for potential errors. Jaccard similarity and Levenshtein Distance were applied to food outlet names and postcode units to calculate a score that reflects the similarity between them. For example, after name standardisation ‘Zizzi – Cambridge’ and ‘Zizzi’ had a Jaccard score of 100 (indication of a perfect score).

We manually reviewed and annotated links in 40 randomly selected postcode sectors to refine matching criteria (Figure 1; n=146,210 rows of potential matches). This annotated data was used to train a Random Forest classifier model (Figure 2), which then matched the remaining unannotated data. Each physical food outlet could only be linked to one online food outlet, and vice versa, ensuring that no duplicate links were made between physical and online outlets. If multiple online food outlets could be linked to the same physical outlet, only one link was included, and the other online food outlets were assumed to be virtual brands.

**Figure 2.**
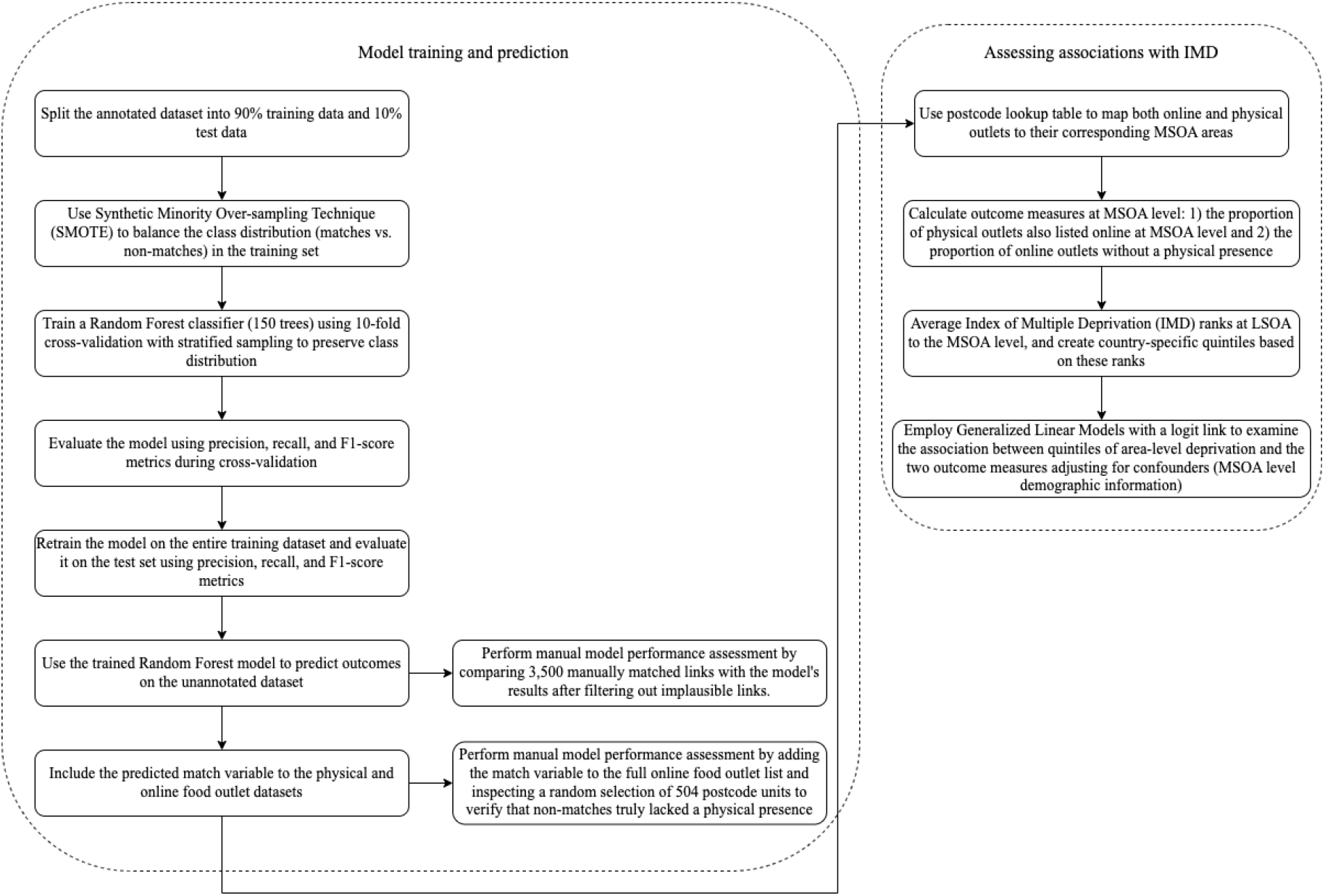
Analytical workflow for matching physical and online food outlets and assessing potential socioeconomic variations

#### Socio-demographic information

Covariates that were hypothesized to influence the association between area-level deprivation and outcome measures were included: population size, proportion of females, proportion of working-age individuals (15-65), and proportion of households with dependents at the MSOA level (for England and Wales) and the intermediate zone level (for Scotland, hereafter also referred to as MSOA) were extracted from 2021 Census data (9, 10).

Indices of Multiple Deprivation (IMD) were obtained from governmental websites at the Lower Super Output Area (LSOA) level for England and Wales (2019), and at the data zone level for Scotland (2020) (11, 12). The IMD ranks of LSOAs and data zones were averaged to determine the IMD rank at the MSOA level, after which country-specific quintiles were developed based on these ranks.

### Statistical analyses

#### Model training and evaluation

The annotated data was split into training (90%) and test (10%) sets (Figure 2). To address the imbalance in the training data the Synthetic Minority Over-sampling Technique (SMOTE) was applied. SMOTE generates synthetic samples of the underrepresented matched outlets, balancing the dataset and preventing the model from being biased toward the more frequent non-matches. This resulted in an equal distribution of matches and non-matches in the training data.

A Random Forest classifier was trained on the training data using 10-fold cross-validation with stratified sampling to maintain class distribution consistency across folds. Cross-validation was employed to generate out-of-sample predictions for the training data, which allowed us to evaluate model performance before retraining it on the full training set. The retrained model was then evaluated on the test set using precision, recall, and F1-score metrics. Finally, the model was applied to the unannotated dataset, following identical data preparation steps, to generate match predictions. Manual inspections of the unannotated dataset were conducted to assess the quality of the matching process. All data preparation and model training were performed using Python.

#### Assessing socioeconomic differences

A lookup table of postcode units was used to map both online and physical food outlets to their corresponding MSOA areas, enabling the calculation of the number of outlets in each MSOA area and the subsequent proportion measures (13).

We used beta regression models to examine the association between quintiles of area-level deprivation and both outcome measures. Marginal effects (at mean) were calculated to interpret the results. All models were adjusted for the covariates listed above and analyses were performed in R version 4.2.3.

## Results

### Matching physical and online food outlets

At the MSOA level, the median number of physical outlets was 32 (IQR 20–53) and the median number of online outlets was 5 (IQR 1–11; Table 1). The cross-validation results on the training set indicated a mean F1 score of 99%, along with a mean recall and precision of 99%. When applied to the test data, the Random Forest model demonstrated class-specific performance metrics with a score of 100% across all three metrics for non-matches. For matches, it yielded an F1 score of 90%, a recall of 98%, and a precision of 83%. The confusion matrix can be found in Supplementary Table 1.

**Table 1.** Food outlet availability characteristics by area-level deprivation at MSOA level in Great Britain (n=8525)

|  | Total<br>(n=8525) | IMD Q1<br>(most<br>deprived)<br>(n=1706) | IMD Q2<br>(n=1705) | IMD Q3<br>(n=1705) | IMD Q4<br>(n=1703) | IMD Q5<br>(least<br>deprived)<br>(n=1706) |
| --- | --- | --- | --- | --- | --- | --- |
| Count of MSOA areas with at least one physical food outlet (%) | 8503 (99.7%) | 1704 (99.9%) | 1701 (99.8%) | 1696 (99.5%) | 1700 (99.8%) | 1702 (99.8%) |
| Count of MSOA areas with at least one online food outlet (%) | 7162 (84.0%) | 1641 (96.2%) | 1508 (88.4%) | 1370 (80.3%) | 1284 (75.4%) | 1359 (79.6%) |
| Median number of physical food outlets (IQR) | 32 (20 – 53) | 36 (21 – 65) | 36 (22 – 60) | 34 (21 – 56) | 32 (20 – 49) | 27 (16 – 40) |
| Median number of online food outlets (IQR) | 5 (1 – 11) | 9 (4 – 17) | 7 (2 – 15) | 4 (1 – 10) | 3 (1 – 7) | 3 (1 – 6) |
| Median percentage of physical outlets also listed online (IQR) | 14% (0 - 23) | 20% (13 - 27) | 18% (8 - 26) | 13% (2 - 23) | 9% (0 - 19) | 9% (2 - 19) |
| Median percentage of online-only outlets (IQR) | 14% (0 - 27) | 17% (0 - 27) | 15% (0 - 25) | 14% (0 - 25) | 13% (0 - 29) | 7% (0 - 27) |

Abbreviations: MSOA = Middle Super Output Area; IMD = Indices of Multiple Deprivation; Q = Quartile; IQR = Interquartile Range
The percentage of physical outlets also listed online was calculated as the number of physical food outlets matched to an online outlet divided by the total number of physical food outlets at MSOA level \* 100. The percentage of online-only outlets was calculated as 1 minus the number of online food outlets matched to a physical outlet divided by the total number of online food outlets at MSOA level \* 100.

The median percentage of physical outlets listed online was 14% (IQR 0-23), while the median for online-only outlets was 14% (IQR 0-27; Table 1). Also, a higher percentage of physical restaurants (median 11%) and takeaways (median 40%) were listed online compared to other categories, such as pubs and supermarkets, which had a median of 0% (Table 2).

**Table 2.** Median percentage of physical outlets also listed online at MSOA level by physical food outlet category as determined by the FSA.

| Physical food outlet category | Median | IQR |
| --- | --- | --- |
| Mobile caterer | 0 | 0 – 0 |
| Other catering premises | 0 | 0 – 0 |
| Pub/bar/nightclub | 0 | 0 – 0 |
| Restaurant/café/canteen | 11 | 0 – 29 |
| Retailers – other | 0 | 0 – 11 |
| Retailers – supermarkets/hypermarkets | 0 | 0 – 33 |
| Takeaway/sandwich shop | 40 | 0 – 59 |

Abbreviations: FSA = Food Standards Agency; IQR = Interquartile Range

### Area-level deprivation differences

The most deprived areas had the greatest median number of physical and online food outlets (Table 1). In the most deprived areas, 20% (IQR 13 - 27) of physical outlets were also listed online, compared to 9% (IQR 2-19) in the least deprived areas. The percentage of online-only outlets was 17% (IQR 0 - 27) in the most deprived areas and 7% (IQR 0 - 27) in the least deprived areas.

We found that the proportion of physical outlets also listed online was lower in less deprived areas (Q3-5 versus Q1; Table 3). For example, lower deprived areas (Q4 and Q5) was associated with a 6% (95%CI 5% - 6%) lower proportion of physical outlets listed online compared to the most deprived area (Q1). Also, the proportion of online-only outlets was lower in less deprived areas (Q3, Q4 and Q5) compared to the most deprived area.

**Table 3.**
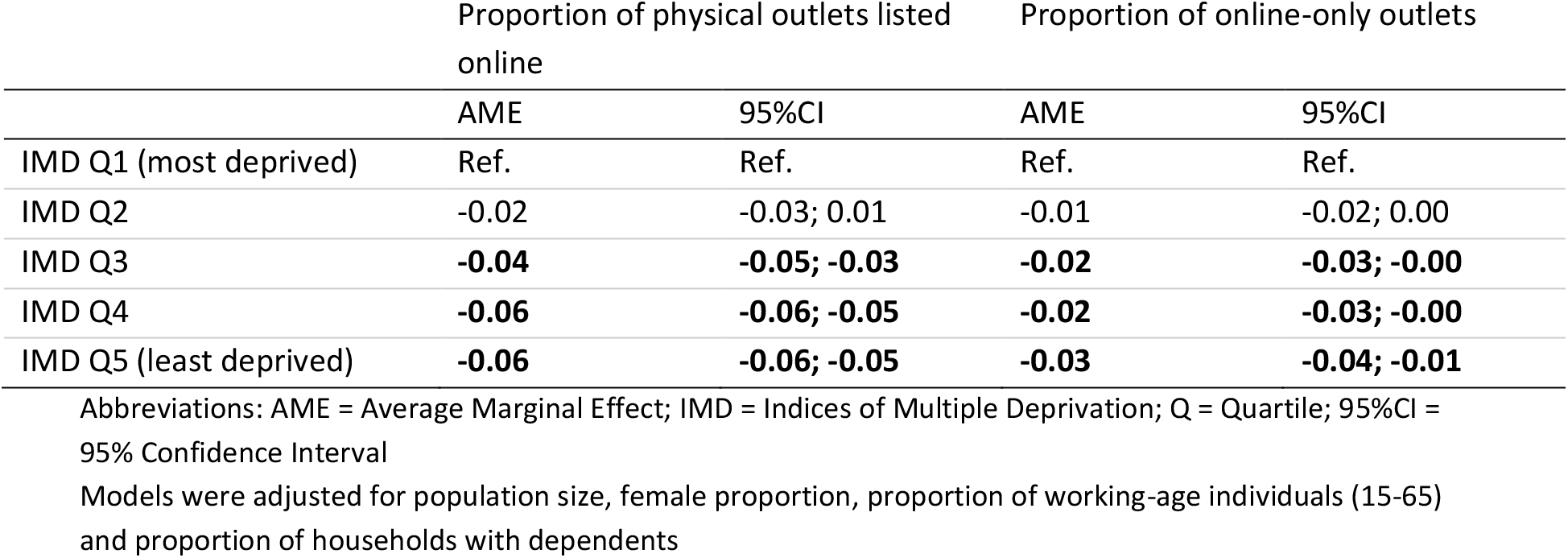
Marginal effects of the association between quartiles of area-level deprivation, and the proportion of physical outlets also listed online (n=8503) and online-only outlets (n=7162)

|  | Proportion of physical outlets listed online |  | Proportion of online-only outlets |  |
| --- | --- | --- | --- | --- |
|  | AME | 95%CI | AME | 95%CI |
| IMD Q1 (most deprived) | Ref. | Ref. | Ref. | Ref. |
| IMD Q2 | -0.02 | -0.03; 0.01 | -0.01 | -0.02; 0.00 |
| IMD Q3 | **-0.04** | **-0.05; -0.03** | **-0.02** | **-0.03; -0.00** |
| IMD Q4 | **-0.06** | **-0.06; -0.05** | **-0.02** | **-0.03; -0.00** |
| IMD Q5 (least deprived) | **-0.06** | **-0.06; -0.05** | **-0.03** | **-0.04; -0.01** |

Abbreviations: AME = Average Marginal Effect; IMD = Indices of Multiple Deprivation; Q = Quartile; 95%CI = 95% Confidence Interval
Models were adjusted for population size, female proportion, proportion of working-age individuals (15-65) and proportion of households with dependents

## Discussion

We matched physical and online food outlets in GB using a Random Forest model, which achieved high cross-validation scores. Analyses revealed that the proportion of physical outlets listed online and online-only outlets was greatest in more deprived areas.

This study demonstrates the efficiency of machine learning for tasks that would otherwise be labour-intensive to perform manually. Our Random Forest model likely reduced human error and made linkages a less tedious task, achieving high F1, precision, and recall scores (83% to 100%). Despite initial concerns about overfitting, the consistent performance across training and test sets—supported by previous research (14)—confirms the model’s reliability. Around 82% of online outlets were successfully matched to physical ones, aligning with our annotated dataset. Additional data cleaning could further improve model performance, as discrepancies were primarily due to name variations (e.g. ‘AFC’ vs. ‘AFC Fried Chicken’) and chain outlets with multiple purposes (e.g. supermarkets with instore food concessions).

We found that the proportion of physical outlets listed online and online-only outlets declined with decreasing deprivation. Previous GB research similarly showed greater availability of online and physical food outlets in more deprived areas (2, 15), which may contribute to unhealthy dietary behaviours, obesity, and socioeconomic differences in health outcomes (16, 17).

A strength of this study was using highly current data with minimal time difference between datasets. The inclusion of a broad range of outlets in the FSA dataset may have affected the results, as focusing on outlets more likely to go online (i.e. takeaways and restaurants) could reduce the number of non-matches and enhance model precision. Additionally, while Just Eat is more widely available and popular, relying on a single OFDS platform may limit generalizability (18). Future research should incorporate additional datasets, such as those from other OFDS platforms. Monitoring the proportion of physical outlets listed online and online-only outlets is important for understanding how the food environment is evolving and for identifying potential impacts on dietary behaviours, health outcomes, and socioeconomic inequalities.

## Conclusion

This study demonstrated the effectiveness of Random Forest models in matching physical food outlets from a centrally held register for food hygiene data to their online counterparts on the most popular UK OFDS platform. The results show that the median percentage of physical outlets with an online presence is 14%, and the median percentage of online-only outlets is also 14%. The current method shows potential for ongoing monitoring of both physical and digital food environments.

## Data Availability

Food Standards Agency data is publicly available. Code to webscrape data from Online Food Delivery Service platforms can be found online.

https://ratings.food.gov.uk/

https://github.com/YuruHuang/Online-food-delivery

## Declarations

### Consent for publication

Not applicable.

### Availability of data and materials

All data used was publicly available. The code used to process the data and run the Random Forest classifier can be found on Github (https://github.com/Jodyyy16/Linking_physical_to_online_FO).

### Authors’ contributions

The authors’ responsibilities were as follows – JCH: study design, conducted analyses and prepared the initial draft of the manuscript; YH: data collection, reviewed and edited the manuscript; JA: study design, supervision, and reviewed and edited the manuscript. All authors read and approved the final manuscript.

## Acknowledgements

We would like to thank Dr Mark Kattenberg at the Netherlands Bureau for Economic Policy Analysis for reviewing the manuscript.

## Declaration of generative AI and AI-assisted technologies in the writing process

During the preparation of this work the author(s) used ChatGPT4o in order to improve readability and language of the manuscript. After using this tool, the authors reviewed and edited the content as needed and take full responsibility for the content of the published article.

## References

1. Hoenink JC, Huang Y, Keeble M, Mackenbach JD, Pinho MG, Burgoine T, et al. Socioeconomic distribution of food outlet availability through online food delivery services in seven European countries: A cross-sectional study. Health & Place. 2023;84:103135.

2. Keeble M, Adams J, Bishop TR, Burgoine T. Socioeconomic inequalities in food outlet access through an online food delivery service in England: A cross-sectional descriptive analysis. Applied Geography. 2021;133:102498.

3. Food Standards Agency. Food Hygiene Rating Scheme [Internet]. Available from: https://www.food.gov.uk/safety-hygiene/food-hygiene-rating-scheme. Accessed September 2024.

4. Office for National Statistics. Web scraping policy [Internet]. Available from: https://www.ons.gov.uk/aboutus/transparencyandgovernance/datastrategy/datapolicies/webscrapingpolicy#:~:text=4.-,Policy%20statement,that%20serve%20the%20public%20good. Accessed September 2024.

5. Wilkins EL, Radley D, Morris MA, Griffiths C. Examining the validity and utility of two secondary sources of food environment data against street audits in England. Nutrition journal. 2017;16:1–13.

6. Statista. Meal Delivery [Internet]. Available from: https://www.statista.com/outlook/emo/online-food-delivery/meal-delivery/united-kingdom?currency=USD&locale=en. Accessed September 2024.

7. Huang Y, Burgoine T, Bishop TR, Adams J. Assessing the healthiness of menus of all out-of-home food outlets and its socioeconomic patterns in Great Britain. Health & Place. 2024;85:103146.

8. YouGov. The most popular dining brands (Q2 2024) [Internet]. Available from: https://yougov.co.uk/ratings/consumer/popularity/dining-brands/all. Accessed September 2024.

9. Nomis. Census 2021 Bulk Download [Internet]. Available from: https://www.nomisweb.co.uk/sources/census_2021_bulk. Accessed September 2024.

10. Statistics Government Scotland. Population Estimates Summary (Current Geographic Boundaries): a data cube spreadsheet [Internet]. Available from: https://statistics.gov.scot/slice?dataset=http%3A%2F%2Fstatistics.gov.scot%2Fdata%2Fpopulation-estimates-2011-datazone-linked-dataset&http%3A%2F%2Fpurl.org%2Flinked-data%2Fsdmx%2F2009%2Fdimension%23refPeriod=http%3A%2F%2Freference.data.gov.uk%2Fid%2Fyear%2F2021&http%3A%2F%2Fstatistics.gov.scot%2Fdef%2Fdimension%2Fage=http%3A%2F%2Fstatistics.gov.scot%2Fdef%2Fconcept%2Fage%2Fall&http%3A%2F%2Fstatistics.gov.scot%2Fdef%2Fdimension%2Fsex=http%3A%2F%2Fstatistics.gov.scot%2Fdef%2Fconcept%2Fsex%2Fall. Accessed September 2024.

11. Gov.uk [Internet]. English indices of deprivation. Available from: https://www.gov.uk/government/collections/english-indices-of-deprivation. Accessed on September 2024.

12. Gov.scot [Internet]. Scottish Index of Multiple Deprivation 2020. Available from: https://www.gov.scot/collections/scottish-index-of-multiple-deprivation-2020/. Accessed on September 2024.

13. Office for National Statistics [Internet]. Postcode to OA (2011) to LSOA to MSOA to LAD (May 2022) Best Fit Lookup in the UK. Available from:. https://geoportal.statistics.gov.uk/datasets/e7824b1475604212a2325cd373946235/about. Accessed on September 2024.

14. Kalbus A, Ballatore A, Cornelsen L, Greener R, Cummins S. Associations between area deprivation and changes in the digital food environment during the COVID-19 pandemic: Longitudinal analysis of three online food delivery platforms. Health & place. 2023;80:102976.

15. Maguire ER, Burgoine T, Monsivais P. Area deprivation and the food environment over time: A repeated cross-sectional study on takeaway outlet density and supermarket presence in Norfolk, UK, 1990–2008. Health & Place. 2015;33:142–7.

16. Burgoine T, Forouhi NG, Griffin SJ, Wareham NJ, Monsivais P. Associations between exposure to takeaway food outlets, takeaway food consumption, and body weight in Cambridgeshire, UK: population based, cross sectional study. Bmj. 2014 Mar 13;348.

17. Burgoine T, Sarkar C, Webster CJ, Monsivais P. Examining the interaction of fast-food outlet exposure and income on diet and obesity: evidence from 51,361 UK Biobank participants. International Journal of Behavioral Nutrition and Physical Activity. 2018 Dec;15:1–2.

18. Hoenink JC, Huang Y, Keeble M, Mackenbach JD, Pinho MGM, Vanderlee L, et al. Physical and online food outlet availability and its influence on dietary behaviours in Great Britain: A repeated cross-sectional study. SSM Pop Health. 2025.

