## Supplemental Table 1 for "Linking physical food outlets to online platforms: A cross-sectional machine learning approach to analysing socioeconomic variations in Great Britain"

Supplementary Table 1. Confusion matrix of the random forest classifier model on the annotated data

| No match true label | 5783 | 23 |
| --- | --- | --- |
| Match true label | 2 | 110 |
|  | No match predicted label | Match predicted label |
